# Clinical profiling of specific diagnostic subgroups of women with chronic pelvic pain

**DOI:** 10.1101/2022.10.03.22280515

**Authors:** Lysia Demetriou, Michal Krassowski, Pedro Abreu Mendes, Kurtis Garbutt, Allison F. Vitonis, Elizabeth Wilkins, Lydia Coxon, Lars Arendt-Nielsen, Qasim Aziz, Judy Birch, Andrew W Horne, Anja Hoffman, Lone Hummelshoj, Claire E Lunde, Jane Meijlink, Danielle Perro, Nilufer Rahmioglu, Kathryn L. Terry, Esther Pogatzki-Zahn, Christine B Sieberg, Rolf-Detlef Treede, Christian M Becker, Francisco Cruz, Stacey A Missmer, Krina T Zondervan, Jens Nagel, Katy Vincent

## Abstract

Chronic pelvic pain (CPP) is a common condition affecting up to 26.6% of women, with many suffering for several years before diagnosis and/or treatment. Its clinical presentation is varied and there are frequently comorbid conditions both within and outside the pelvis. We aim to explore whether specific subgroups of women with CPP report different clinical symptoms and differing impact of pain on their quality of life (QoL).

The study is part of the Translational Research in Pelvic Pain (TRiPP) project which is a cross-sectional observational cohort study. The study includes 769 female participants of reproductive age who completed an extensive set of questions derived from standardised WERF EPHect questionnaires. Within this population we defined a control group (reporting no pelvic pain, no bladder pain syndrome, and no endometriosis diagnosis, N=230) and four pain groups: endometriosis-associated pain (EAP, N=237), interstitial cystitis/bladder pain syndrome (BPS, N=72), comorbid endometriosis-associated pain and BPS (EABP, N=120), and pelvic pain only (PP, N=127).

Clinical profiles of women with CPP (13-50 years old) show variability of clinical symptoms. The EAP and EABP groups scored higher than the PP group (p<0.001) on the pain intensity scales for non-cyclical pelvic pain and higher than both the BPS and PP groups (p<0.001) on the dysmenorrhoea scale. The EABP group also had significantly higher scores for dyspareunia (p<0.001), even though more than 50% of sexually active participants in each pain group reported interrupting and/or avoiding sexual intercourse due to pain in the last 12 months.

Scores for the QoL questionnaire (SF-36) reveal that CPP patients had significantly lower QoL across all SF-36 subscales (p<0.001). Significant effects were also observed between the pain groups for pain interference with their work (p<0.001) and daily lives (p<0.001), with the EABP suffering more compared to the EAP and PP groups (p<0.001).

Our results demonstrate the negative impact that chronic pain has on CPP patients’ QoL and reveal an increased negative impact of pain on the comorbid EABP group. Furthermore, it demonstrates the importance of dyspareunia in women with CPP. Overall, our results demonstrate the need for further exploration of interventions targeting QoL more broadly and suggest that novel approaches to classifying women with CPP are needed.

## Introduction

Chronic pelvic pain (CPP) is common, affecting up to 26.6% of women (1–4), yet it remains difficult to treat. For some women an associated pathology such as endometriosis, adenomyosis, inflammatory bowel disease or an entrapment neuropathy can be identified, but for many others diagnostic investigations will be normal and the label chronic pelvic pain syndrome (CPPS) applied. However, even in those women where, for example, endometriosis is identified the lack of correlation between measures of disease severity and pain symptoms (5,6) makes it challenging to understand the extent to which symptoms are actually caused by endometriosis and to predict the benefit of treatments targeting the ectopic tissue.

There is increasing evidence that chronic pain conditions share many features whatever the underlying cause (7,8), leading some to suggest that chronic pain should be treated as a condition in itself (9,10). Whilst there is relatively limited research on this in the context of chronic pelvic pain, studies that have looked at these features confirm similarities with other chronic pain conditions such as irritable bowel syndrome (IBS), rheumatoid arthritis and fibromyalgia (11,12). However, current clinical practice guidelines are still relatively focussed on the identification and treatment of underlying pelvic pathology (13).

The Translational Research in Pelvic Pain (TRiPP) project (https://www.imi-paincare.eu/PROJECT/TRIPP/) aims to better understand the mechanisms generating and maintaining pelvic pain with a particular focus on endometriosis and interstitial cystitis/bladder pain syndrome (IC/BPS) (14). Here, we use the baseline data from the TRiPP project to determine whether women with CPP in different diagnostic groups report different clinical symptoms and impact on their lives or whether in fact these diagnostic labels do not help us stratify this patient group. A better understanding of the relationship between clinical symptoms, diagnostic grouping and quality of life (QoL) would allow the development of refined clinical pathways and the prioritisation of specific subgroups of women for further research. Given the diversity of symptoms described by women with all forms of CPP we do not expect to see differences in specific clinical symptoms between those with and without endometriosis or with and without IC/BPS (with the exception of bladder symptoms themselves). However, we hypothesise that those women with comorbid pain and bladder symptoms will describe a poorer quality of life.

## Methods

### Ethical Approval

The study has received ethical approval from Yorkshire & The Humber – South Yorkshire Research Ethics Committee (19/YH/0030) with local site approvals and was being conducted in accordance with the principles of the Declaration of Helsinki and with relevant regulations and Good Clinical Practice. Informed consent was obtained from all the participants, and they were informed that they are free to withdraw their data from the study at any time.

### Study population

As described in the TRiPP protocol paper (14), participants were identified from two existing endometriosis cohort studies in Oxford (EndOX: A study to identify possible biomarkers in women with endometriosis, Oxford REC ref 09/H0604/58) (N=276) and Boston (The Women’s Health Study from Adolescence to Adulthood (A2A), IRB-P00004267) (N=494) plus 16 BPS participants who were recruited at Hospital São João/Instituto de Biologia Molecular e Celular (IBMC) in Porto (Supplementary I). It is important to note that both parent studies also included control participants with no pelvic pain symptoms.

The study comprised of five study groups and participants were assigned according to specific inclusion and exclusion criteria for each study group (14) (Table I): endometriosis-associated pain (EAP: N=237), comorbid endometriosis associated- and bladder pain syndrome (EABP: N= 120), bladder pain syndrome (BPS: N=72), pelvic pain only (PP: N=127) and controls (CON: N=230)). Information relating to these criteria was available in the baseline questionnaires participants completed when recruited to the parent study in UOXF and BCH. At IBMC participants were clinically diagnosed with BPS and it was reconfirmed after completion of the baseline questionnaire that they met the criteria for inclusion in this group.

**Table I.**
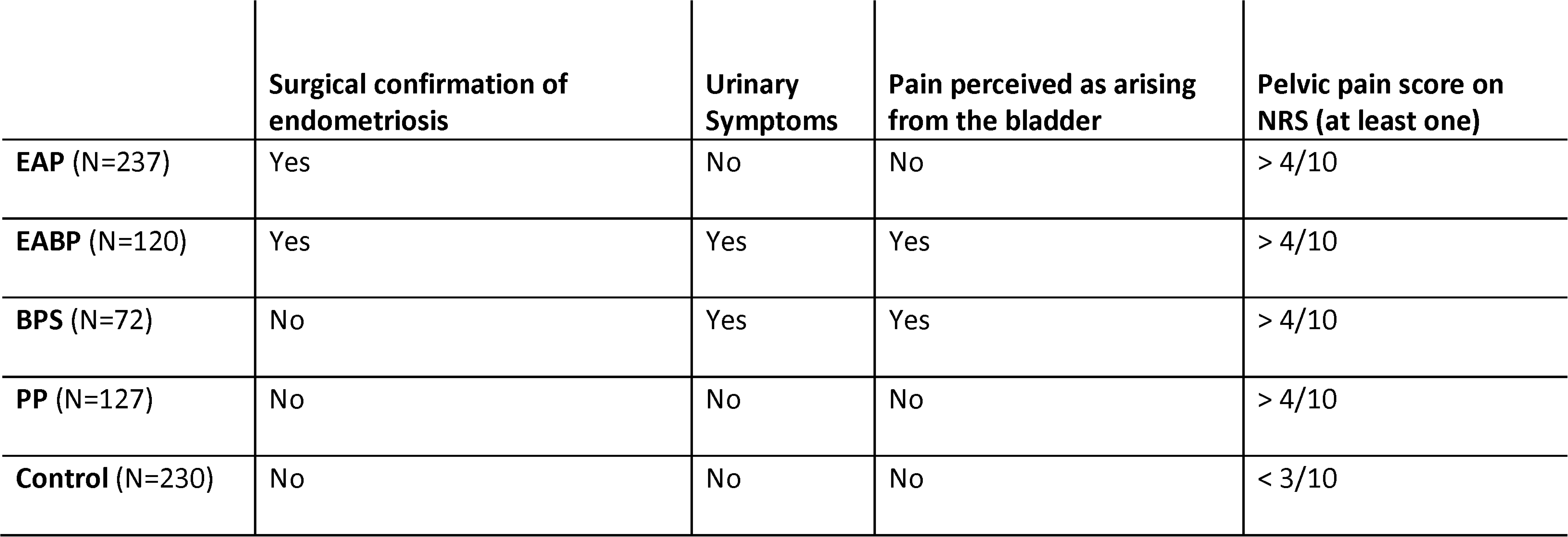
Study design and inclusion criteria for the five study groups: endometriosis-associated pain (EAP, endometriosis-associated d bladder pain (EABP), bladder pain syndrome (BPS), pelvic pain only (PP) and controls).

All participants were women of reproductive age (13-50 years). Pregnant, lactating or women planning pregnancy during the course of the study were not included as per the study exclusion criteria.

### Study design

The TRiPP study design is described in Supplementary I (and more fully elsewhere (14)), the present report focuses on Phase I in which participants completed expanded World Endometriosis Research Foundation (WERF) Biobanking Harmonisation Project (EPHect) questionnaires (15). Here we explore a selected set of outcomes as described below that will also be used in the analysis of phases II and III of the TRiPP project.

### Study Data

#### Demographics and Reproductive History

Demographic information collected included participants’ age, body mass index (BMI), race, education, work and relationship status. Information about the participants’ gynaecological history consisted of: occurrence and frequency of pelvic pain, menstrual history (e.g. menarche and dysmenorrhoea onset), use of hormones in the last 3 months and information on fertility and dyspareunia. Data from a number of questions were analysed to give a fuller picture of sexual activity, associated pain and functional impact. The questions on dyspareunia were not included in the first version of the questionnaire that was completed by 67 participants in Oxford and for participants younger than 18 years old (N=79) from the Boston cohort.

#### Pelvic pain

Pain was assessed using Numerical Rating Scales (NRS) for intensity ranging from 0 = “no pain” to 10 = “worst imaginable pain”. Participants were asked to complete NRS for the experience of pelvic pain at its “worst” in the last three months (for non-cyclical pelvic pain), during the last period (for dysmenorrhoea), worst bladder pain in the last 7 days and pain during and 24hours post sexual intercourse (for dyspareunia).

The Pain Catastrophising Scale (PCS) (16) was used to assess pain-related worry. Catastrophizing is the single most important risk factor that impairs the effectiveness of pain-relieving interventions (17,18), and it statistically mediates the prospective influence of factors such as anxiety on pain outcomes (19). The PCS is a validated/standardised 13-item questionnaire that uses a 5-point scale ranging from 0 (not at all) to 4 (all the time), the total PCS score was computed by summing the responses to all 13 items (16). The scores for the three subscales: Rumination (e.g.,“I can’t stop thinking about how much it hurts.”),, Magnification (e.g., “I worry that something serious may happen.”) and Helplessness (e.g., “There is nothing I can do to reduce the intensity of my pain.”) were computed by summing responses of specific items as outlined in Sullivan (16). Scores above the “clinical cut-off points” were considered as clinically relevant levels of catastrophising (PCS Total: 30, Rumination: 11, Magnification: 5, Helplessness: 13).

#### Medical comorbidities

Participants were provided with a list of medical conditions and reported whether they had received a medical diagnosis for any of those at any point in their lives. The list included autoimmune, gynaecological, mental health, chronic pain, endocrine and cardiovascular disorders.

Furthermore, given the prevalence of bowel symptoms in association with both endometriosis and IC/BPS, irritable bowel syndrome (IBS) was assessed based on the Rome III criteria using a question about the participants’ bowel movements/stools when they experienced non-cyclical pelvic pain in the last 3 months. The symptoms consisted of: a) pelvic pain getting worse/better after bowel movement, b) more/less frequent bowel movement when pain started and c) looser/harder stools when the pain started. Participants who reported having two or more bowel symptoms for “most of the time” or “always” met the criteria for IBS.

#### Quality of life

The participants’ health status was assessed using the Short Form Health Survey (SF-36) which consists of 36 questions covering eight health domains: physical functioning, physical role limitations, bodily pain, general health, emotional role limitations and mental health. The questionnaire was scored as per instructions by Rand Corporation (https://www.rand.org/health-care/surveys_tools/mos/36-item-short-form/scoring.html) and Burholt et al. (20). A higher score indicates a better health status.

To assess pain interference with the participants’ lives we used a question assessing the impact of pain on six different aspects (work or school, daily activities at home, sleep, sexual intercourse, exercise/sports and social activities). Participants had to rate the extent of pain interference using a 4-point Likert scale ranging from “Not at all” to “Extremely” for the last 3 months. The scores were recoded into four groups: low, medium, high and no pain interference or not applicable. Additionally, two NRSs were used to assess the severity of the impact of pelvic pain on the participants’ work and personal daily activities productivity during the past 4 weeks.

#### Factors worsening and relieving pain

Two multiple-choice questions were used to assess/identify factors that participants believe worsen or relieve their pelvic pain. Among others, the options included visceral functions such as bowel movement or bladder emptying; behavioural including exercising, standing/walking or sitting; wellbeing factors like stress and meditation; or other factors e.g. time of day and having a full meal.

### Statistical analysis

The data were entered into a REDCap database (21) and analysed using IBM SPSS Statistics software, version 27. The analysis was conducted in 2 phases. First, descriptive analysis was run to identify the characteristics of each study group. Then, statistical testing was undertaken to investigate any effects between the study groups for the study outcomes. Statistical testing for the NRS pelvic and bladder pain did not include the control group since they were recruited based on their NRS scores being <3/10. One-way ANOVAs and Bonferroni corrected post-hoc tests were used to compare between study groups for all the continuous variables while frequencies, percentages and non-parametric tests (chi-square, Mann-Whitney U tests) were employed for all the categorical variables. Pearson’s correlations were run to explore relationships between the variables. Bonferroni correction was used to account for multiple comparisons.

## Results

### Demographics and Reproductive history

A total of 785 participants were included in this phase of the study with a mean age of 27.6 years (STD: 8.1) and mean BMI of 24.8 (STD: 5.2) (Table II). A one-way ANOVA showed significant differences for age but not for BMI between groups (Age: F(4,781)=4.12, p=0.003, BMI: F(4, 757)=3.28, p=0.011); post-hoc independent-samples t-tests showed that the BPS group was significantly older than the EABP (p=0.01) and Controls (p=0.04).

**Table II.**
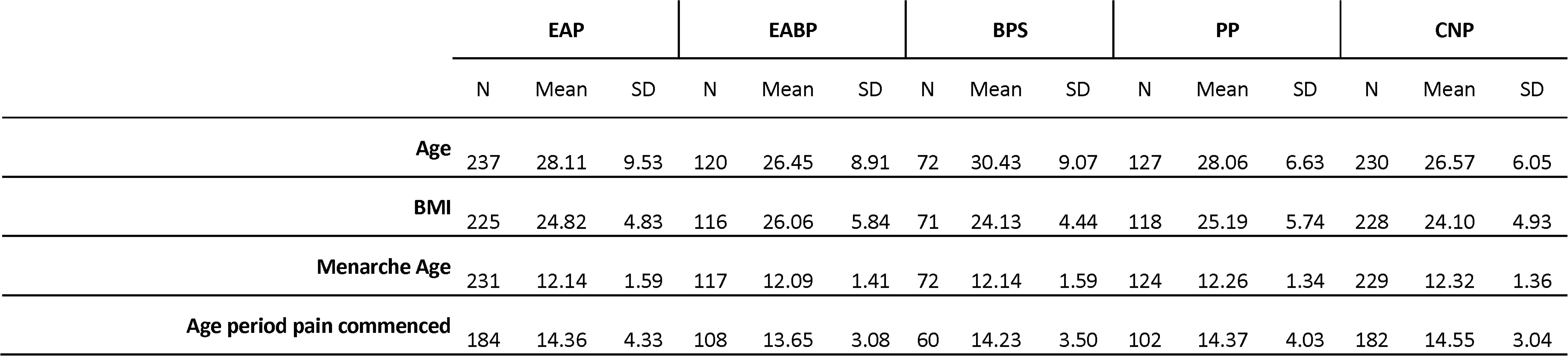
Key participant characteristics (age, body mass index (BMI), age of menarche and age period pain commenced). Data shown as numbers (N), ean and standard deviation (SD).

The age of menarche (mean=12.2 years) ranged from 8 to 16 years old (One-way ANOVA: F(4,768)=0.715, p=0.582) and did not significantly differ among groups (Table II). Likewise, the age at which dysmenorrhoea commenced was similar among all study groups (mean=14.3 years, F(4,631)=1.10, p=0.356) (Table II). On average, patients suffered for approximately 8 years with non-cyclical pelvic pain, and there were no significant differences between the patient groups, even though the PP group had the lowest mean number of years (mean=4.6 years), while the EABP and BPS groups had the highest mean number of years (EABP: mean=9.7 years, BPS: mean=9.9 years) (F(4,204)=3.906, p=0.004). A Pearson’s correlation showed that the number of years suffering with pelvic pain positively correlated with the age of the participants (r=0.411, p<0.001).

More participants in the PP (90.5%) and Controls (92.6%) group reported having periods during the last three months than participants in the EAP (76.9%), EABP (63.9%) and BPS (71.8%) (Table III). Of the participants who reported having periods, more than 53.8% of the controls and 35.0% of the pain groups reported having periods whilst using exogenous hormones in the last 3 months. The most common reason for not having periods across the groups was taking hormones continuously (91.8%). Regardless of whether they had periods, many participants reported using exogenous hormones in the last three months (EAP: 52.7%, EABP: 61.7%, BPS: 61.1%, PP: 39.4%, Controls: 57%) (Table III).

**Table III.**
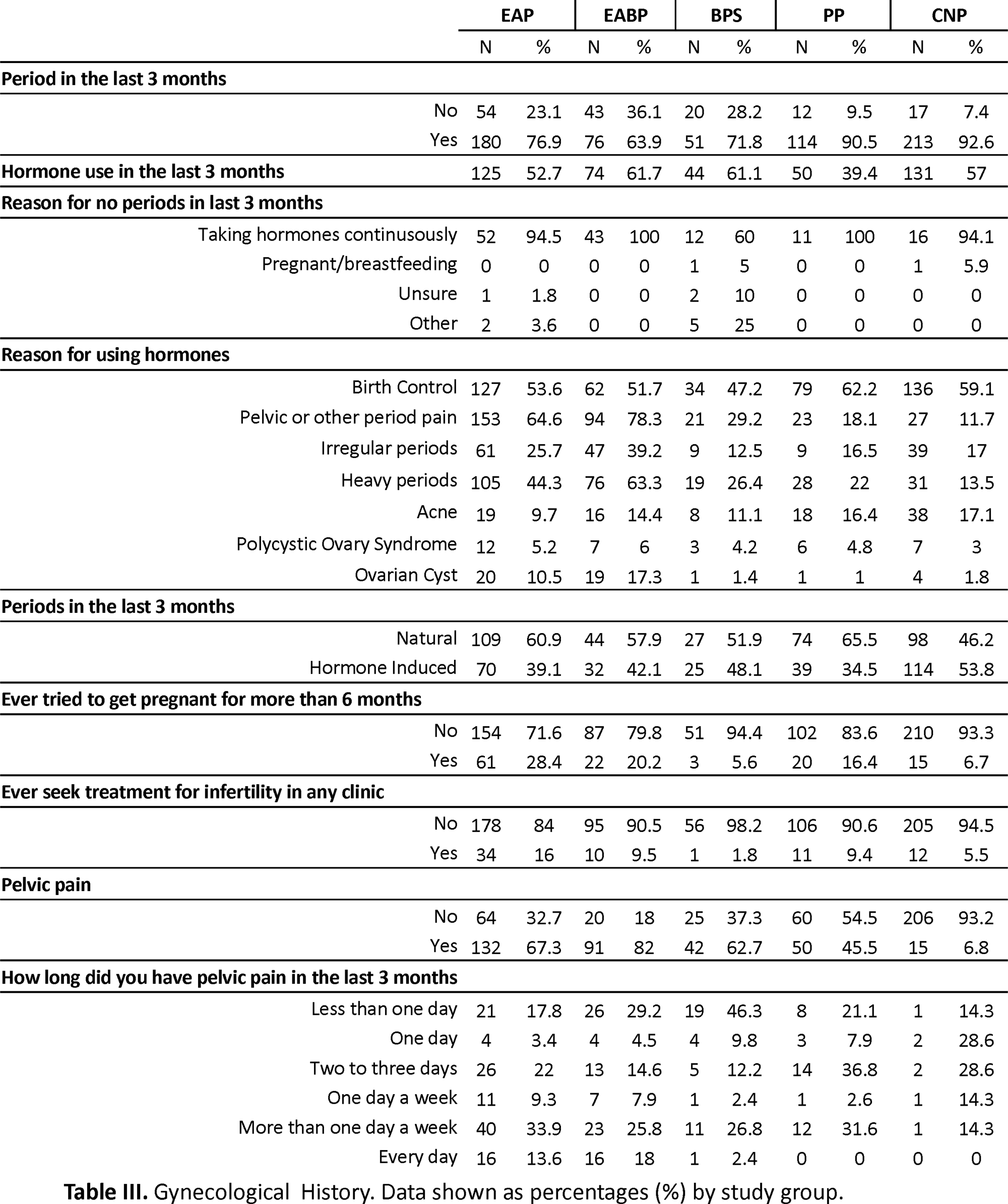
Gynecological History. Data shown as percentages (%) by study group.

### Pelvic pain

The pain rating for non-cyclical pain at its worst during the last three months was significantly higher in the EAP and EABP compared to the PP group (Table IV). When asked about dysmenorrhoea at its worst the EABP and EAP groups had a significantly higher mean score on pain severity compared to both the BPS and PP groups (p<0.001). Results for the NRS scale on bladder pain during the last seven days confirmed significantly higher mean pain scores for the BPS and EABP groups compared to the EAP and PP groups (p<0.001) as defined by the inclusion criteria (Figure I).

**Table IV.**
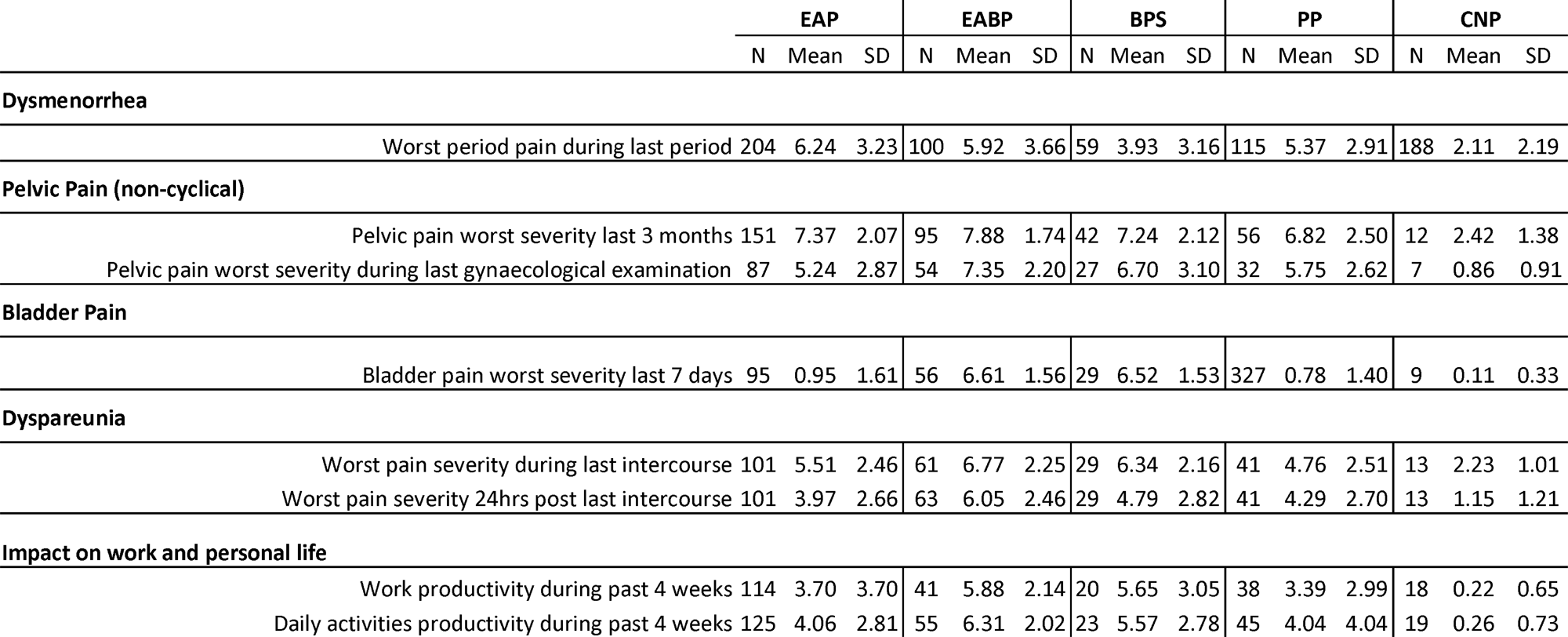
Mean score of the Numerical Rating Scales (NRS) on dysmenorrhea, non-cyclical pelvic pain, bladder pain, dyspareunia and impact of on work and personal life. Data shown as numbers (N), mean and standard deviation (SD).

**Figure 1.**
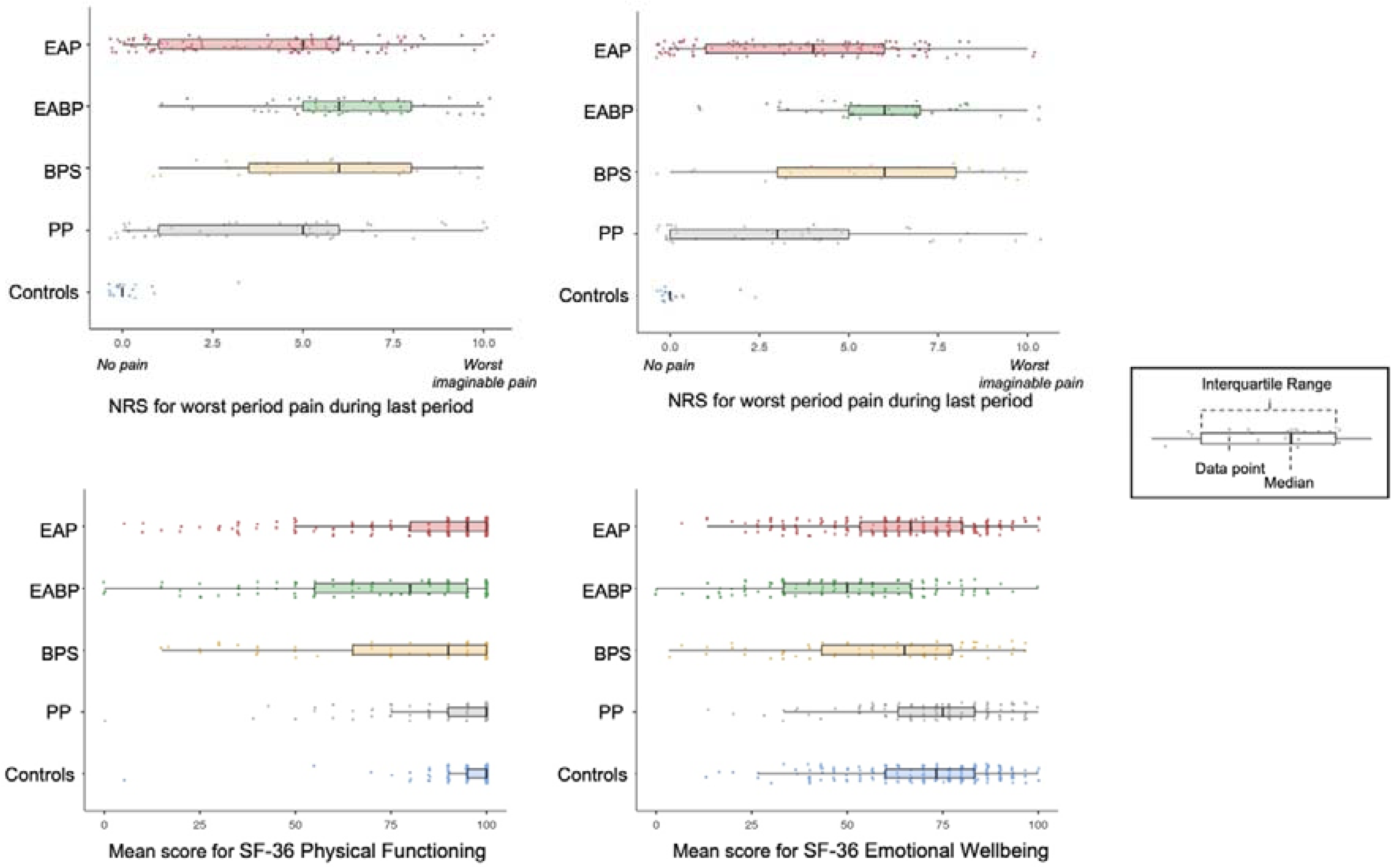
Mean scores and distribution of the Numerical Rating Scales (NRS) for the five study groups on dysmenorrhea (EAP: N=204, EBAP: N=100, BPS: N=59, PP: N=115), non-cyclical pelvic pain (EAP: N=151, EBAP: N=95, BPS: N=42, PP: N=56), dyspareunia (EAP: N=101, EABP: N=61, BPS: N=29, PP: N=41) and post-coital pain (EAP: N=101, EBAP: N=63, BPS: N=29, PP: N=41).

Regarding questions on dyspareunia, there were missing responses for more than 50% of our participants, reasons for missingness include: a) 4.8% (n=38) of all the participants did not wish to respond to the questions, b) 7.5% (n=59) reported never having sexual intercourse c) these questions were not available to 10.1% (n=79) due to their age at time of completion as described in the methods section above and d) 20.2% (n=150) reported not having sex in the last 12 months and therefore were only included in the analysis focussed on ever having experienced pain with or after intercourse.

From the available data sexual intercourse data however, most participants in each of the pain groups (>58%) reported that they have experienced pain during intercourse or in the 24 hours following vaginal sexual intercourse (Table V). Of those, more than 60% experienced dyspareunia during the last month across all the pain groups. Almost half of those participants (49.2%) in the EABP group said that they had always experienced dyspareunia during intercourse in the last 12 months as opposed to the BPS and PP groups in which most participants reported occasional dyspareunia in the last year. In the EAP group >77% of participants reported experiencing dyspareunia often, usually or always (Table V).

**Table V.**
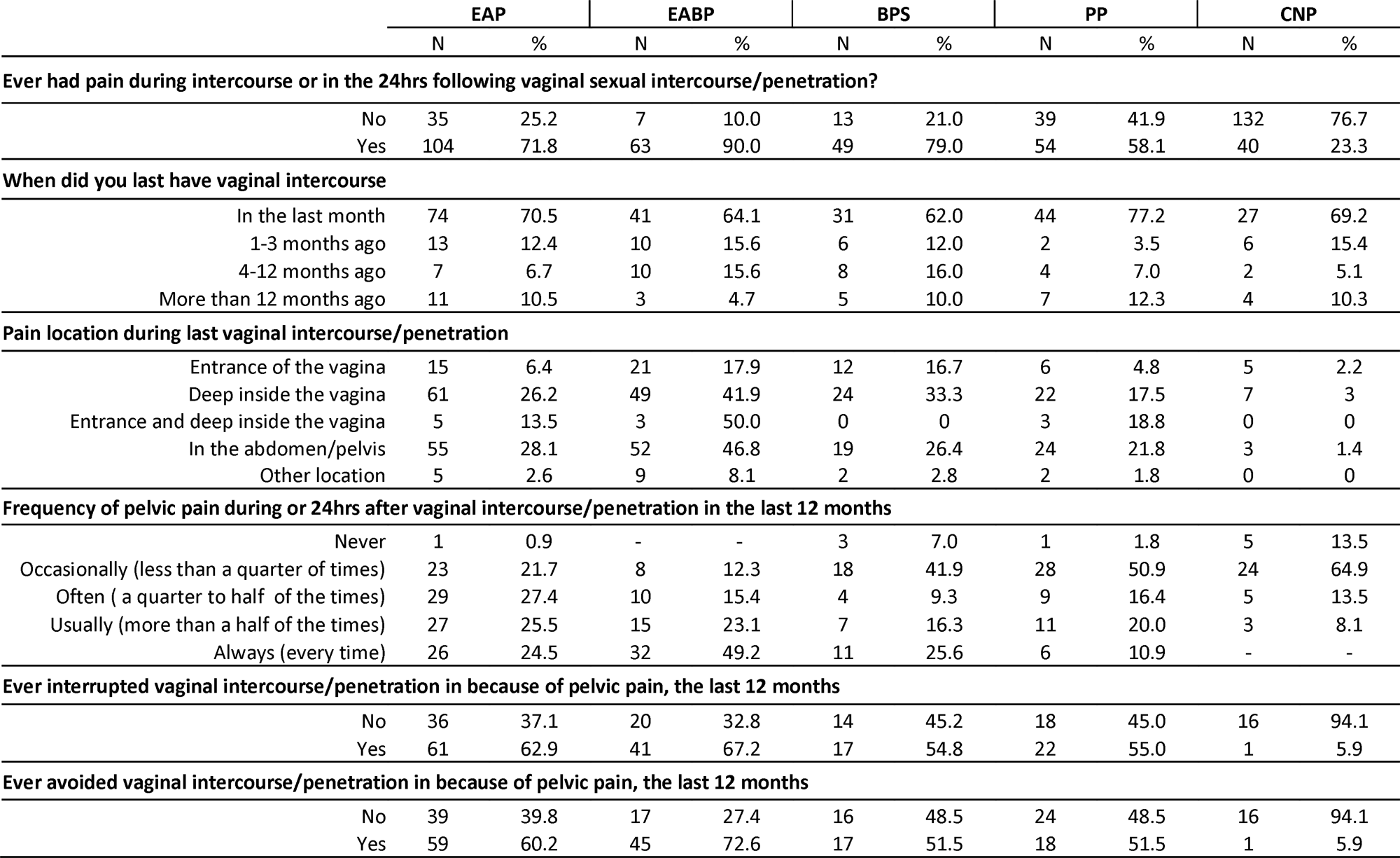
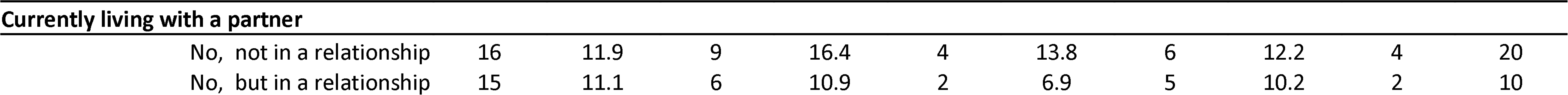
Mean score of the Numerical Rating Scales (NRS) on dysmenorrhea, non-cyclical pelvic pain, bladder pain, dyspareunia and impact of on work and personal life. Data shown as numbers (N), mean and standard deviation (SD).

Concerning the location of dyspareunia, most participants in each pain group reported feeling pain deep inside the vagina, the second most common location was in the abdomen/pelvis. When asked whether they have been avoiding or interrupting sexual intercourse during the last 12 months due to pelvic pain, more than half of the participants (>54%) in all four pain groups said that they have interrupted sexual intercourse while more than half (>51%) of the participants in the EAP, EABP and BPS groups reported avoiding it.

Further analysis showed effects between the groups for the two dyspareunia NRS scales (pain during intercourse: F(4,240)=12.77, p<0.001, pain in the 24hrs post-intercourse: F(4,242)=12.33, p<0.001) (Figure I). The EABP group had significantly higher mean ratings on the NRS scales than all other pain groups (p<0.001) for the pain felt during intercourse, and significantly higher mean rating for pain severity in the 24hours after the sexual intercourse than the EAP and PP groups (Table IV and Figure I).

### Medical comorbidities

Across all pain groups the most common comorbidities were depression, anxiety, migraine, asthma, and irritable bowel syndrome (IBS) (Table VI). Within the EAP, EABP and BPS groups there were higher percentages of depression, anxiety and migraine diagnosis (>28%) than in the PP group (<22%), while the frequency of asthma ranged between 20% - 27% across all pain groups (Table VI). Statistical analysis using Mann-Whitney U tests revealed that when compared to the control group only the EABP group has significantly higher frequency of anxiety diagnosis (anxiety: (U(N_EABP_=114, N_CON_=221)=10510, z=-3.39, p<0.001), and depression: (U(N_EABP_=120, N_CON_=230)=10855, z=-4.36, p<0.001)). However, statistical analysis for migraine showed that it was significantly more common in EAP (28.7%), EABP (34.2%) and BPS (40.2%) groups than the control group (9.6%) ((U(N_EAP_=237, N_CON_=230)=22042, z=-5.23, p<0.001), (U(N_EABP_=120, N_CON_=230)=10405, z=-5.68, p<0.001), (U(N_BPS_=72, N_CON_=230)=5737, z=-6.60, p<0.001)) groups. Similarly, IBS diagnosis was also significantly more common in the EAP (18.6%), EABP (24.2%) and BPS (20.8%) groups when compared to the control group (4.8%) ((U(N_EAP_=237, N_CON_=229)=23402, z=-4.60, p<0.001), (U(N_EABP_=120, NCON=230)=11079, z=-5.39, p<0.001), (U(NBPS=72, NCON=229)=6922, z=-4.22, p<0.001)).

**Table VI.**
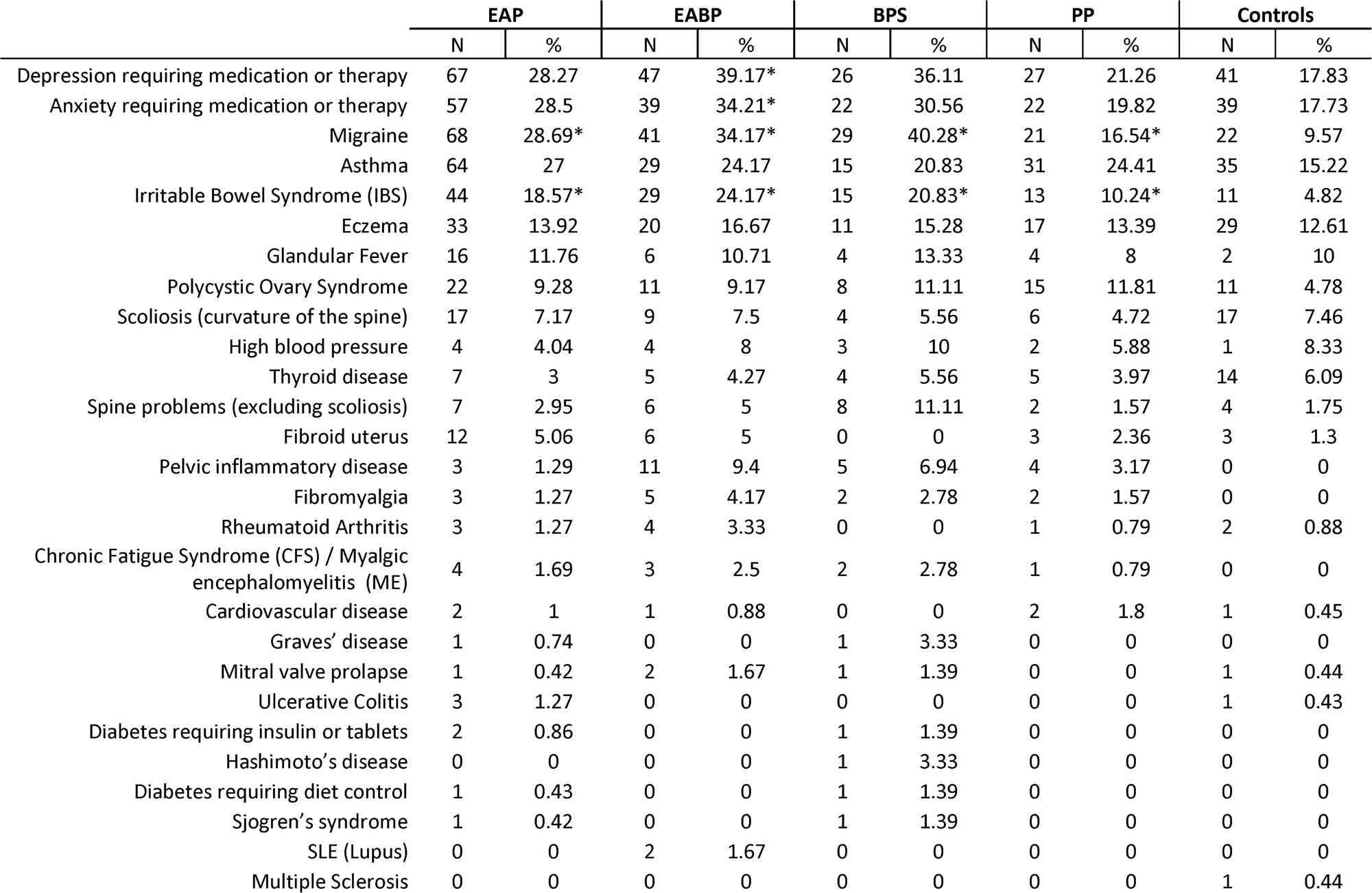
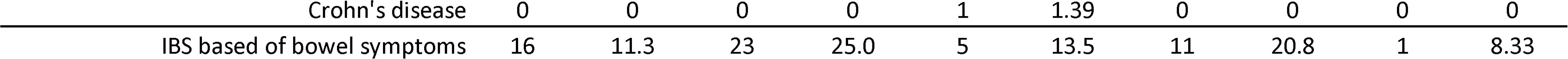
Comorbidities occurrence in our population. Data shown as numbers (N) and percentages (%). * represents significant difference between ain groups and the control group (p<0.001) using a Mann-Whitney U test.

IBS diagnosis was also less common in the PP group (10%) compared to the EAP, EABP and BPS groups (>18%).

Further analysis of the bowel symptoms revealed that more than 11% of the participants in the pain groups met the criteria for IBS regardless of whether they reported having a medical diagnosis for it in the comorbidities question. The EABP and PP groups had the higher percentages of participants meeting the IBS criteria (>20%) (Table VI).

### Quality of life

Analysis of the SF-36 questionnaire using a one-way ANOVA revealed significant effects between the five study groups across all eight SF-36 domains (p<0.001). Specifically, the EABP group suffered the most while the control group had the best scores across all sub-scales (p<0.001) (Table VII). Significant effects were also observed between the four pain groups for pain interference with activities at work and daily life (work: F(3,209)=9.76, p<0.001; daily activities: F(3,244)=10.51, p<0.001) (Figure II). Post-hoc tests revealed that the EABP group had significantly higher pain interference scores than the EAP and PP groups at both scales (p<0.001) (Table IV).

**Table VII.**
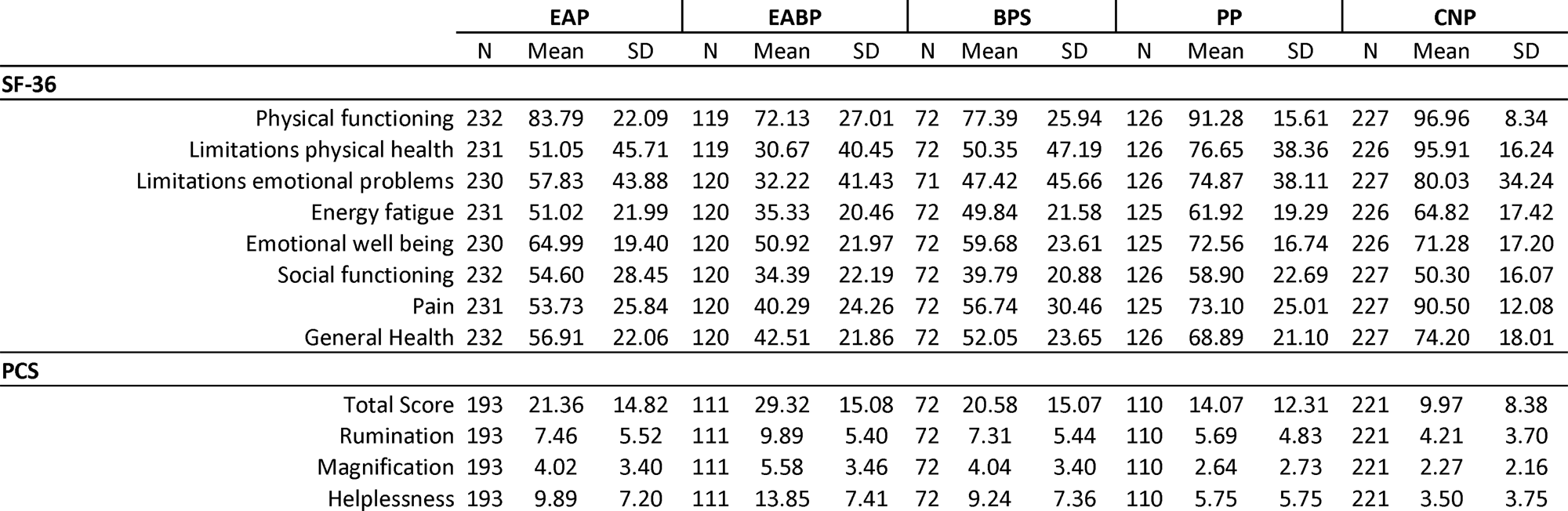
Quality of life SF-36 and Pain Catastrophising Questionnaire (PCS) scores by study group. Data shown as numbers (N), mean and standard eviation (SD).

**Figure 2.**
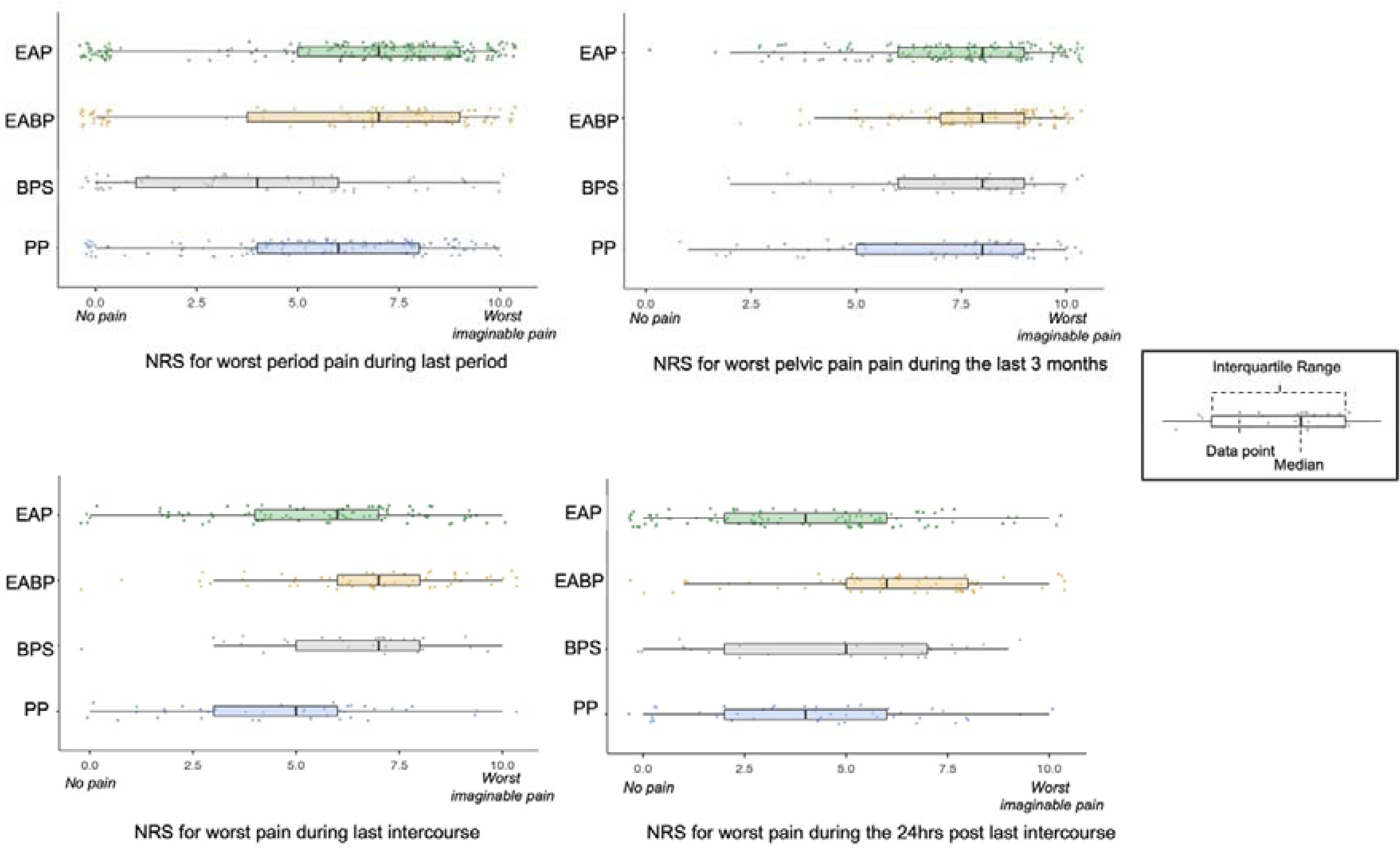
Mean scores and distribution of the Numerical Rating Scales (NRS) (impact of pain on work (EAP: N=114, EBAP: N=41, BPS: N=20, PP: N=38, CNP: N=18) and personal life (EAP: N=125, EBAP: N=55, BPS: N=23, PP: N=45, CNP: N=19)); and the physical functioning and emotional wellbeing sub-scales of the SF-36 questionnaire (physical functioning (EAP: N=232, EBAP: N=119, BPS: N=72, PP: N=126, CNP: N=227) and emotional wellbeing (EAP: N=230, EBAP: N=120, BPS: N=72, PP: N=125, CNP: N=226)) for the five study groups.

### Pain Catastrophising

Pain catastrophising was higher in the EABP group both in the mean total score and in the mean scores of all three subscales (p<0.001). The EAP and BPS groups also had a higher pain catastrophising score than both the PP and Controls groups (p<0.001). According to the clinical-cut-offs more than 19% of the participants in the EAP, EABP and BPS had clinical levels of pain catastrophising. The EABP group had the highest percentage of people above the clinical cut-off (36.0%) (Table VII). A Pearson’s correlation between all the pain catastrophising and all the NRS pain scales across all CPP patients revealed strong positive correlations (p<0.001).

### Factors worsening and relieving pain

On average, participants from all groups chose three to four factors that either relieve or worsen their pelvic pain. Across the four pain groups the three most common factors for worsening pelvic pain were: stress (23.6%), full bladder/urinating (23.3%) and exercising (20.2%). The most frequently reported factors for relieving pelvic pain were: pain medication (31.4%), lying down (31.0%) and heating pad (29.5%) (Table IX). Analysis of bowel movement and bladder emptying as visceral factors, shows that less people in the EAP and the PP groups report these factors as relieving (bowel movement: EAP: 19.8% and PP 15.7%; emptying bladder: EAP: 9.3% and PP 4.7%) or worsening (bowel movement: EAP: 6.7% and PP 2.5%; emptying bladder: EAP: 24.1% and PP 10.2%), while in the EABP and BPS groups more participants report visceral factors as worsening (bowel movement: EABP: 27.7% and BPS 9.5%; emptying bladder: EABP: 65.8% and BPS 44.4%) rather than relieving (bowel movement: EABP: 32.5% and BPS 20.8%; emptying bladder: EAP: 34.2% and BPS 23.6%).

**Table VIII.**
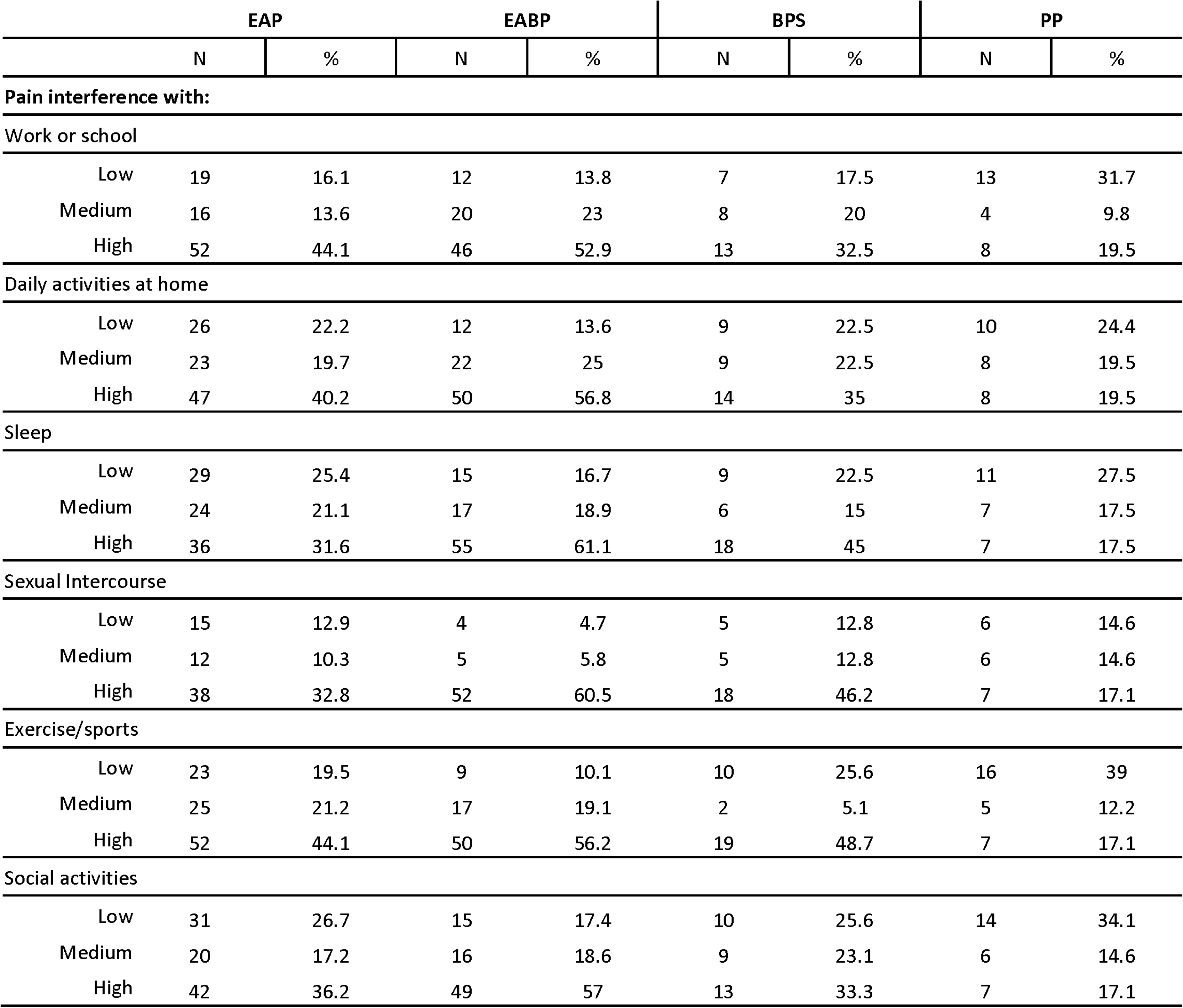
Information for pain interference on work or school, daily activities at home, sleep, sexual life, exercise or sports and social activities. Data shown as numbers (N), mean and standard deviation (SD).

**Table IX.**
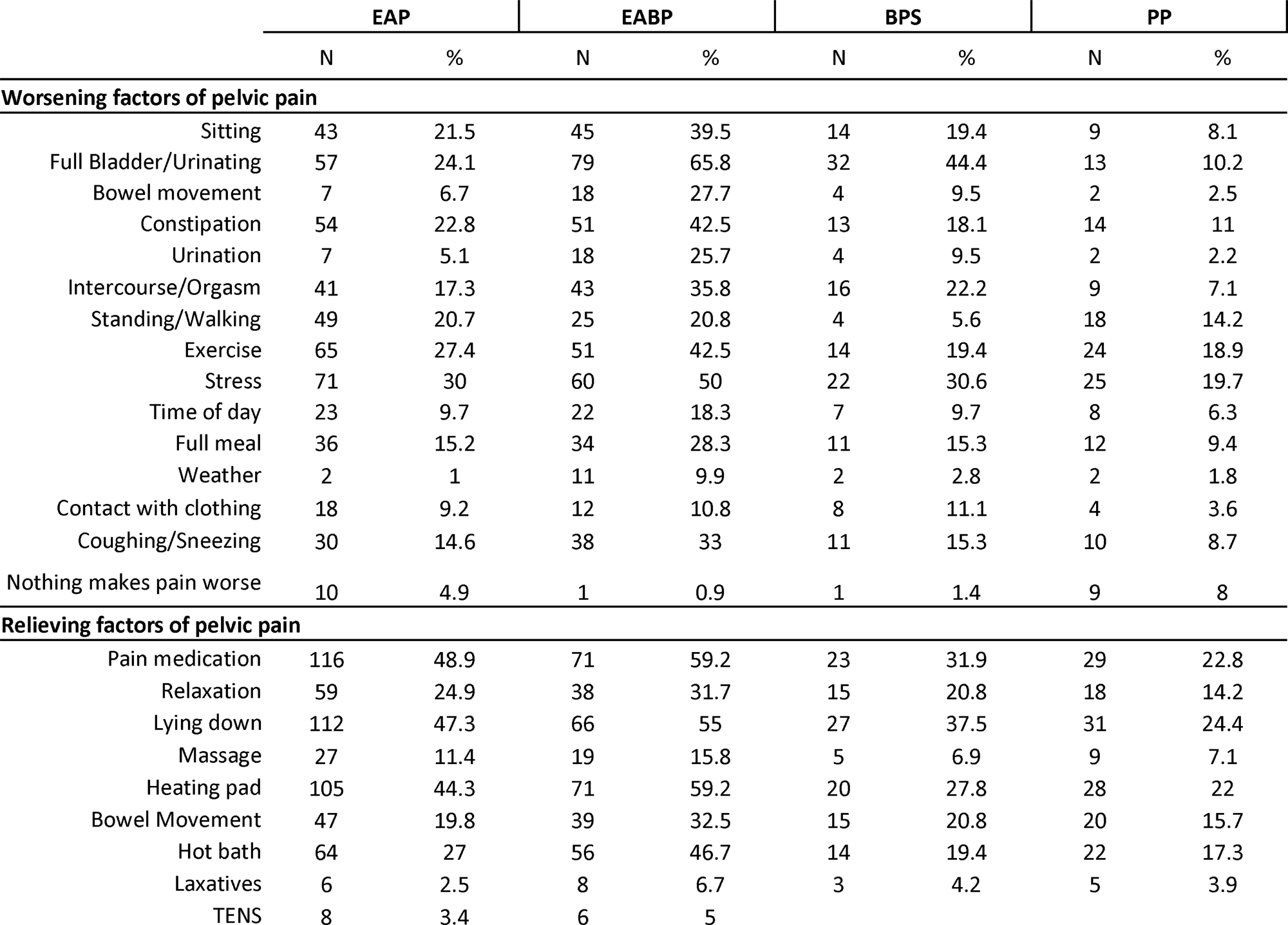
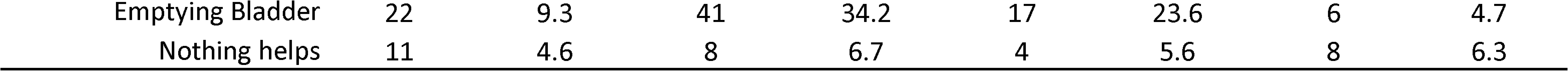
Worsening and relieving factors for pelvic pain by study group. Data shown as numbers (N), mean and standard deviation (SD).

## Discussion

This study phenotypically describes the clinical profiles of women with CPP, with a particular focus on endometriosis and IC/BPS, and contrasts them with controls. In line with existing literature, we have demonstrated that regardless of the aetiology, CPP has a negative impact on the lives of people who suffer with it. However, our results show that there are important differences between different clinical subgroups of those with CPP and those with comorbid endometriosis and bladder pain syndrome (EABP) are particularly severely affected. The heterogeneity in factors exacerbating and relieving pelvic pain, highlights the complexity of the condition but suggests that there may be a variety of different underlying pain mechanisms contributing to the similar clinical presentation. Further evaluation of these underlying mechanisms may provide more clinically relevant approaches to patient stratification than focussing on diagnosis alone.

Importantly, we highlight how common painful sexual intercourse is in women with CPP and that half of those who answered these questions reported interrupting or avoiding it due to pain. This is an important issue for women and their partners and it can have a negative effect on more than one aspect of a woman’s life, including mental health, body image, relationship with their partner and fertility (22,23). Dyspareunia is a commonly described symptom of endometriosis, but there is less of a focus on this issue in IC/BPS. It is therefore particularly notable that the highest dyspareunia scores were reported by those with bladder symptoms in our cohort. This is in line with previous work in women with endometriosis, demonstrating that the presence of tenderness of the bladder/pelvic floor or a diagnosis of IC/BPS is associated with more severe dyspareunia in those with all stages of endometriosis (24). A better understanding of the mechanisms underlying dyspareunia in all women with CPP not just those with deep endometriosis is clearly an important future need and will ultimately improve the clinical care of those suffering with this important symptom (25).

It is perhaps unsurprising that those with comorbid endometriosis and bladder pain symptoms scored highly on NRSs for all types of pelvic pain (non-cyclical pelvic pain, bladder pain, dysmenorrhoea and dyspareunia). However, our data demonstrate that these women also have poorer quality of life and report greater pain interference. Whilst there are studies exploring the prevalence and comorbidity of endometriosis and subsequent diagnosis of IC/BPS (26–28), few studies focus on those with comorbid symptoms, either in terms of exploring clinical features or considering specific treatment regimes. Moreover, for many clinical studies the presence of a comorbidity is an exclusion criteria and thus very limited data exists, this is particularly true for clinical trials. Our findings suggest this may be a priority group to consider in future work.

Our study focuses on two specific diagnoses, endometriosis and IC/BPS, however there are many other diagnoses associated with CPP. The EPHect Clinical Covariates questionnaire captures some of these: IBS, fibroids, pelvic inflammatory disease, inflammatory bowel disease and a limited number of musculoskeletal conditions (spine problems including scoliosis, rheumatoid arthritis and fibromyalgia). However, for most of these the numbers were relatively small within our cohort and therefore it is not possible to explore their contribution to the clinical picture in any detail. There are also a number of other factors that have been associated with chronic pain more broadly and with CPP specifically, including psychological and social factors, adverse childhood events and other traumatic experiences (13). The data collected within the EPHect questionnaire does not allow these to be taken into consideration here. Additional information is collected on some of these measures for a smaller cohort in Phase II of TRiPP and will be interrogated to better understand their relationship to the pain experience and mechanisms underlying any observed relationship (14).

Given that viscero-visceral referral is a relatively well understood phenomenon (29) and that visceral pain conditions are often comorbid (30), we had expected to see higher rates of IBS in those with bladder symptoms. This was the case when questioning previous diagnosis of IBS (EABP 24% and BPS 21% vs EAP 19% and PP 10%). However, considering the answers to the questions comprising the Rome IV criteria, a higher proportion of those in the EABP and PP groups would be considered to meet IBS criteria than in the other groups. It is of course difficult to know whether this reflects ongoing effective treatment in at least a proportion of those with a past diagnosis of IBS. Nonetheless, the high rates of IBS across the cohort as a whole highlight the importance of assessing symptoms from all abdominopelvic organs in women with CPP.

Interestingly the impact of visceral function (bladder and bowel emptying specifically) on pelvic pain appears to be very variable. For some participants these visceral functions worsened their pain, whilst for others they could be relieving. In fact, there was wide variability in the factors identified as either worsening or relieving pelvic pain even within diagnostic groups and it would be interesting to consider whether patterns within these data may reflect underlying pain mechanisms rather than for example the presence or absence of endometriosis or bladder symptoms. Whilst it can feel challenging in a clinical consultation to raise the topics of mental health and the stressors of daily life, particularly when women may have had a long journey to get their pain taken seriously, stress was the most commonly identified worsening factor whereas relaxation was one of the most frequent relieving factors. Moreover, anxiety and depression were commonly reported comorbidities and those in the EAP, EABP and BPS groups all scored significantly lower than controls on the emotional function dimension on the quality of life questionnaire. This evidence suggests that patients could benefit from treatments focusing on managing their stress and developing better coping mechanisms for their pelvic pain.

Whilst not an assessment of psychological wellbeing, catastrophic thinking about pain has been shown to be an important predictor of outcomes both in the transition to chronic pain after an acute injury and in the response to a variety of therapeutic options (31,32). In line with other studies (33,34) we observed that the chronic pain groups (EAP, EABP and BPS) scored higher in pain catastrophising compared to the control group with a particularly high percentage of EABP patients meeting clinical levels of catastrophising. This highlights the need to include behavioural interventions that specifically target pain-related worry in the management of CPP in the same way as they are considered essential to the management of other chronic pain conditions (35,36).

### Strengths and weakness

There are a number of strengths to the TRiPP cohort which include the size of the sample and the detailed phenotypic information available for these participants. Moreover, our study focuses on chronic pelvic pain and therefore includes women with a variety of underlying diagnoses rather than studying only a single condition such as endometriosis. This strategy allows us to compare the similarities between different clinical subgroups but also to understand differences between them. We have a particular interest in IC/BPS and therefore this cohort is relatively unusual in allowing the comparison between endometriosis and IC/BPS as well as those with comorbid symptoms. Unfortunately, our BPS group is smaller than we initially planned. Neither of the original cohorts from which the TRiPP sample was formed had specifically recruited IC/BPS patients (although many did meet the diagnostic criteria) and therefore we had planned to expand these with a large number of new recruits from specialist centres. However, the COVID-19 pandemic halted our ability to recruit to the study. Overall, we still consider our sample size sufficient to draw meaningful conclusions (192 participants with bladder pain, 120 of whom have coexisting endometriosis).

A further strength of our study is that participants were recruited from three different centres rather than a single site. However, the duration of recruitment to the original cohorts has meant that the baseline questionnaires have been reviewed and updated such that not all participants completed the same version. This has reduced the number of variables we could analyse across the full cohort. Nonetheless the EPHect questionnaire is very comprehensive and the data we do have available allows us to phenotypically characterise our participants in a very detailed manner.

### Conclusion

Overall, our results demonstrate the negative impact that chronic pain has on the lives of those with CPP and particularly the importance of dyspareunia. Whilst we do see similarities across the specific clinical subgroups that we explored our data highlight the difference between these groups and importantly illustrate how severely impacted the group with comorbid endometriosis and IC/BPS are. The heterogeneity seen in many of our measures, especially the factors exacerbating and relieving pain suggests that multiple different mechanisms may underlie the similar clinical presentations and emphasise the importance of better characterising those with CPP to identify clinically meaningful methods of patient stratification. Further studies within the TRiPP project will explore these mechanisms in greater detail towards this aim.

## Supporting information

Reasons for using exogenous hormones. Data shown as percentages (%) by study group.

Mean score for the Numerical Rating Scales (NRS) for each study groups by site of data collection.

Key participant characteristics for each study groups by site of data collection.

Naturally or hormonally induced periods in the last 3 months presented by study group and by site of data collection.

Assessment-tools employed at each study phase based on the domains of interest.

## Data Availability

All data produced in the present study are available upon reasonable request to the authors

## Conflict of interest

LD, MK, EW, LC, DP, NR, AFV, LAN, QA, JB, AH, LH, CEL, JM, CS, PAM, KG: declare no competing interests

AWH: : reports grant funding from the MRC, NIHR, CSO, Wellbeing of Women, Roche Diagnostics, Astra Zeneca, Ferring, Charles Wolfson Charitable Trust and Standard Life. His employer has received consultancy fees from Roche Diagnostics, AbbVie, Nordic Pharma and Ferring, outside the submitted work. In addition, AWH has a patent for a serum biomarker for endometriosis pending.

AH: Employee of Bayer AG, Germany

EPZ: received financial support from Grunenthal and Mundipharma for research activities and advisory and lecture fees from Grünenthal, Novartis and Mundipharma. In addition, she receives scientific support from the German Research Foundation (DFG), the Federal Ministry of Education and Research (BMBF), the German Federal Joint Committee (G-BA) and the Innovative Medicines Initiative (IMI) 2 Joint Undertaking under grant agreement No 777500. This Joint Undertaking receives support from the European Union’s Horizon 2020 research and innovation programme and EFPIA. All money went to the institution E.M.P.-Z. is working for.

RDT: Ad board for BAYER, IASP task force on chronic pain classification

CMB: Research Grants from Bayer Healthcare, MDNA Life Sciences, Roche Diagnostics, European Commission, NIH. His employer has received consulancy fees from Myovant and ObsEva for work outside of this project

FC: Consultant and/or investigator for Allergan (Abbvie), Astellas, Bayer, Ipsen and Recordati

SAM: has been an advisory board member for AbbVie and Roche and receives research funding from the National Institutes of Health, the US Department of Defense, the J. Willard and Alice S. Marriott Foundation, and AbbVie; none are related to the presented work; the J. Willard and Alice S. Marriott Foundation supported enrollment of and data collection from the A2A cohort in Boston from which TRiPP data were sampled.

KTZ: reports grant funding from EU Horizon 2020, NIH US, Wellbeing of Women, Bayer AG, Roche Diagnostics, Evotec-Lab282, MDNA Life Sciences, outside the submitted work. JN: Employee and shareholder of Bayer AG, Germany

KV: declares research funding from Bayer Healthcare and UKRI and honoraria for consultancy and talks and associated travel expenses from Bayer Healthcare, Grunenthal GmBH, AbbVie and Eli Lilly.

## Author contributions

LD and KV drafted the manuscript. All other authors contributed to the original design of the project and reviewed and edited the manuscript.

## Funding

The study was funded by Innovative Medicines Initiative 2 Joint Undertaking (JU) under grant agreement No 777500. The JU receives support from the European Union’s Horizon 2020 research and innovation programme and EFPIA. Financial support was provided by the J. Willard and Alice S. Marriott Foundation for establishment of and baseline data collection within the A2A cohort - from which the Boston-based TRiPP population was sampled.

## Notes

### Competing Interest Statement

LD, MK, EW, LC, DP, NR, AVF, LAN, QA, JB, AH, LH, CEL, JM, CS, PAM, KG: declare no competing interests
AWH: : reports grant funding from the MRC, NIHR, CSO, Wellbeing of Women, Roche Diagnostics, Astra Zeneca, Ferring, Charles Wolfson Charitable Trust and Standard Life. His employer has received consultancy fees from Roche Diagnostics, AbbVie, Nordic Pharma and Ferring, outside the submitted work. In addition, AWH has a patent for a serum biomarker for endometriosis pending.
AH: Employee of Bayer AG, Germany
EPZ: received financial support from Grunenthal and Mundipharma for research activities and advisory and lecture fees from Grunenthal, Novartis and Mundipharma. In addition, she receives scientific support from the German Research Foundation (DFG), the Federal Ministry of Education and Research (BMBF), the German Federal Joint Committee (G-BA) and the Innovative Medicines Initiative (IMI) 2 Joint Undertaking under grant agreement No 777500. This Joint Undertaking receives support from the European Union's Horizon 2020 research and innovation programme and EFPIA. All money went to the institution E.M.P.-Z. is working for.
RDT: Ad board for BAYER, IASP task force on chronic pain classification
CMB: Research Grants from Bayer Healthcare, MDNA Life Sciences, Roche Diagnostics, European Commission, NIH. His employer has received consulancy fees from Myovant and ObsEva for work outside of this project
FC: Consultant and/or investigator for Allergan (Abbvie), Astellas, Bayer, Ipsen and Recordati
SAM: has been an advisory board member for AbbVie and Roche and receives research funding from the National Institutes of Health, the US Department of Defense, the J. Willard and Alice S. Marriott Foundation, and AbbVie; none are related to the presented work; the J. Willard and Alice S. Marriott Foundation supported enrollment of and data collection from the A2A cohort in Boston from which TRiPP data were sampled.
KTZ: reports grant funding from EU Horizon 2020, NIH US, Wellbeing of Women, Bayer AG, Roche Diagnostics, Evotec-Lab282, MDNA Life Sciences, outside the submitted work.
JN: Employee and shareholder of Bayer AG, Germany
KV: declares research funding from Bayer Healthcare and UKRI and honoraria for consultancy and talks and associated travel expenses from Bayer Healthcare, Grunenthal GmBH, AbbVie and Eli Lilly.

### Clinical Protocols

https://www.medrxiv.org/content/medrxiv/early/2022/05/16/2022.05.16.22274828.full.pdf

### Funding Statement

This study was funded by the Innovative Medicines Initiative 2 Joint Undertaking under grant agreement No 777500. This Joint Undertaking receives support from the European Union Horizon 2020 research and innovation programme and EFPIA Companies. Financial support was provided by the J. Willard and Alice S. Marriott Foundation for establishment of and baseline data collection within the A2A cohort from which the Boston-based TRiPP population was sampled.

### Author Declarations

Ethics committee of the Yorkshire & The Humber South Yorkshire Research Ethics Committee gave ethical approval for this work.

### Summary of Updates

This version of the manuscript has been revised after a peer-review that it has undergone at the Reproductive Health Frontiers Journal.

