## Supplementary material for "Clinical profiling of specific diagnostic subgroups of women with chronic pelvic pain": Reasons for using exogenous hormones. Data shown as percentages (%) by study group.

|  | **EAP (%)** | **EABP (%)** | **BPS (%)** | **PP (%)** | **Controls (%)** |
| --- | --- | --- | --- | --- | --- |
| **Birth control** | 53.6 | 51.7 | 47.2 | 62.2 | 59.1 |
| **Pelvic or other period pain** | 64.6 | 78.3 | 29.2 | 18.1 | 11.7 |
| **Irregular periods** | 25.7 | 39.2 | 12.5 | 16.5 | 17 |
| **Heavy periods** | 44.3 | 63.3 | 26.4 | 22 | 13.5 |
| **Acne** | 9.7 | 14.4 | 11.1 | 16.4 | 17.1 |
| **Polycystic Ovary Syndrome** | 5.2 | 6 | 4.2 | 4.8 | 3 |
| **Ovarian Cyst** | 10.5 | 17.3 | 1.4 | 1 | 1.8 |

**Supplemental Table I.** Reasons for using exogenous hormones. Data shown as percentages (%) by study group.
