## Supplementary material for "Clinical profiling of specific diagnostic subgroups of women with chronic pelvic pain": Mean score for the Numerical Rating Scales (NRS) for each study groups by site of data collection.

|  | **EAP** | | | | | | **EABP** | | | | | | **BPS** | | | | | | | | | **PP** | | | | | | **CNP** | | | | | |
| --- | --- | --- | --- | --- | --- | --- | --- | --- | --- | --- | --- | --- | --- | --- | --- | --- | --- | --- | --- | --- | --- | --- | --- | --- | --- | --- | --- | --- | --- | --- | --- | --- | --- |
|  | **Oxford** | | | **Boston** | | | **Oxford** | | | **Boston** | | | **Oxford** | | | **Boston** | | | **Porto** | | | **Oxford** | | | **Boston** | | | **Oxford** | | | **Boston** | | |
|  | **N** | **Mean** | **SD** | **N** | **Mean** | **SD** | **N** | **Mean** | **SD** | **N** | **Mean** | **SD** | **N** | **Mean** | **SD** | **N** | **Mean** | **SD** | **N** | **Mean** | **SD** | **N** | **Mean** | **SD** | **N** | **Mean** | **SD** | **N** | **Mean** | **SD** | **N** | **Mean** | **SD** |
| **Dysmenorrhea** | | | | | | | | | | | | | | | | | | | | | | | | |  |  |  |  |  |  |  |  |  |
| Worst period pain during last period | 136 | 7.36 | 2.01 | 101 | 4.98 | 3.84 | 56 | 8.02 | 1.62 | 64 | 4.52 | 3.96 | 14 | 8 | 1.33 | 42 | 2.85 | 2.59 | 16 | 4.22 | 3.35 | 50 | 6.86 | 2.04 | 77 | 4.44 | 2.99 | 20 | 2.33 | 1.44 | 210 | 2.09 | 2.23 |
| **Pelvic Pain (non-cyclical)** | | | | | | | | | | | | | | | | | | | | | | | | |  |  |  |  |  |  |  |  |  |
| Pelvic pain worst severity last 3 months | 136 | 7.21 | 2.15 | 101 | 7.64 | 1.9 | 56 | 8.13 | 1.39 | 64 | 7.55 | 2.1 | 14 | 8.85 | 1.07 | 42 | 5.31 | 2.14 | 16 | 7.5 | 1.46 | 50 | 7.47 | 2.18 | 77 | 5.65 | 2.66 | 20 | 2.5 | 1.97 | 210 | 2.33 | 0.52 |
| Pelvic pain worst severity during last gynaecological examination | 136 | 5.24 | 2.87 |  |  |  | 56 | 7.35 | 2.2 |  |  |  | 14 | 8.67 | 1.37 |  |  |  | 16 | 5.13 | 3.23 | 50 | 5.75 | 2.63 |  |  |  | 20 | 0.86 | 0.9 |  |  |  |
| **Bladder Pain** | | | | | | | | | | | | | | | | | | | | | | | | |  |  |  |  |  |  |  |  |  |
| Bladder pain worst severity last 7 days | 136 | 0.95 | 1.61 |  |  |  | 56 | 6.61 | 1.56 |  |  |  | 14 | 6.36 | 0.84 |  |  |  | 16 | 6.67 | 1.99 | 50 | 0.78 | 1.4 |  |  |  | 20 | 0.11 | 0.33 |  |  |  |
| **Dyspareunia** | | | | | | | | | | | | | | | | | | | | | | | | |  |  |  |  |  |  |  |  |  |
| Worst pain severity during last intercourse | 136 | 5.44 | 2.63 | 101 | 5.69 | 1.93 | 56 | 6.67 | 2.3 | 64 | 7.06 | 2.14 | 14 | 7.56 | 1.24 | 42 | 5.25 | 2.34 | 16 | 6.63 | 2.07 | 50 | 4.97 | 2.61 | 77 | 4.25 | 2.26 | 20 | 1 | 1 | 210 | 2.6 | 0.7 |
| Worst pain severity 24hrs post last intercourse | 136 | 3.69 | 2.7 | 101 | 4.77 | 2.41 | 56 | 6.07 | 2.39 | 64 | 6 | 2.72 | 14 | 6.89 | 1.17 | 42 | 4 | 3.22 | 16 | 3.62 | 2.39 | 50 | 4.45 | 2.76 | 77 | 3.92 | 2.64 | 20 | 0 | 0 | 210 | 1.5 | 1.18 |

**Supplemental Table II.** Mean score of the Numerical Rating Scales (NRS) on dysmenorrhea, non-cyclical pelvic pain, bladder pain, dyspareunia and impact of pain on work and personal life for the study groups by site of data collection (Oxford, Boston, Porto). Data shown as numbers (N), mean and standard deviation (SD).
