## Supplementary material for "Clinical profiling of specific diagnostic subgroups of women with chronic pelvic pain": Key participant characteristics for each study groups by site of data collection.

|  | **EAP** | | | | | | **EABP** | | | | | | **BPS** | | | | | | | | | | **PP** | | | | | | | **CNP** | | | | | | |
| --- | --- | --- | --- | --- | --- | --- | --- | --- | --- | --- | --- | --- | --- | --- | --- | --- | --- | --- | --- | --- | --- | --- | --- | --- | --- | --- | --- | --- | --- | --- | --- | --- | --- | --- | --- | --- |
|  | **Oxford** | | | **Boston** | | | **Oxford** | | | **Boston** | | | **Oxford** | | | **Boston** | | | **Porto** | | | | **Oxford** | | | **Boston** | | | | **Oxford** | | | | **Boston** | | |
|  | **N** | **Mean** | **SD** | **N** | **Mean** | **SD** | **N** | **Mean** | **SD** | **N** | **Mean** | **SD** | **N** | **Mean** | **SD** | **N** | **Mean** | **SD** | **N** | **Mean** | **SD** | **N** | | **Mean** | **SD** | **N** | **Mean** | **SD** | **N** | | **Mean** | **SD** | **N** | | **Mean** | **SD** |
| **Age** | 136 | 34.09 | 7.21 | 101 | 20.06 | 5.48 | 56 | 32.38 | 7.39 | 64 | 21.27 | 6.61 | 14 | 30.86 | 7.16 | 42 | 25.48 | 4.26 | 16 | 43.06 | 7.5 | 50 | | 31.64 | 6.81 | 77 | 25.73 | 5.39 | 20 | | 34.15 | 7.47 | 210 | | 25.85 | 5.39 |
| **BMI** | 136 | 25.1 | 4.63 | 101 | 24.47 | 5.06 | 56 | 26.8 | 5.88 | 64 | 25.46 | 5.79 | 14 | 25.63 | 3.77 | 42 | 23.83 | 5.11 | 16 | 23.7 | 2.65 | 50 | | 26.88 | 4.67 | 77 | 24.28 | 6.08 | 20 | | 26.79 | 6.97 | 210 | | 23.86 | 4.67 |
| **Menarche Age** | 136 | 12.48 | 1.62 | 101 | 11.7 | 1.44 | 56 | 12.58 | 1.35 | 64 | 11.69 | 1.34 | 14 | 12.36 | 1.78 | 42 | 12.12 | 1.52 | 16 | 12 | 1.67 | 50 | | 12.26 | 1.63 | 77 | 12.26 | 1.14 | 20 | | 12.58 | 1.64 | 210 | | 12.3 | 1.33 |
| **Age period pain commenced** | 136 | 15.81 | 5.5 | 101 | 12.98 | 2.02 | 56 | 14.44 | 3.06 | 64 | 13.08 | 2.99 | 14 | 13.75 | 2.83 | 42 | 14.5 | 3.86 | 16 | 13.63 | 2.56 | 50 | | 15.1 | 5.63 | 77 | 14.08 | 3.19 | 20 | | 13.17 | 1.17 | 210 | | 14.6 | 3.08 |

**Supplemental Table III.** Key participant characteristics (age, body mass index (BMI), age of menarche and age period pain commenced) for the study groups by site of data collection (Oxford, Boston, Porto). Data shown as numbers (N), mean and standard deviation (SD).
