## Supplementary material for "Clinical profiling of specific diagnostic subgroups of women with chronic pelvic pain": Naturally or hormonally induced periods in the last 3 months presented by study group and by site of data collection.

|  | **EAP** | | | | **EABP** | | | | **BPS** | | | | | | **PP** | | | | **CNP** | | | |
| --- | --- | --- | --- | --- | --- | --- | --- | --- | --- | --- | --- | --- | --- | --- | --- | --- | --- | --- | --- | --- | --- | --- |
|  | **Oxford** |  | **Boston** |  | **Oxford** |  | **Boston** |  | **Oxford** |  | **Boston** |  | **Porto** |  | **Oxford** |  | **Boston** |  | **Oxford** |  | **Boston** |  |
|  | **N** | **%** | **N** | **%** | **N** | **%** | **N** | **%** | **N** | **%** | **N** | **%** | **N** | **%** | **N** | **%** | **N** | **%** | **N** | **%** | **N** | **%** |
| **Periods in the last 3 months** | | | | | | | | | | | | | | | | | | | | | | |
| **Natural** | 88 | 79.28 | 21 | 30.88 | 25 | 69.44 | 19 | 47.5 | 7 | 70 | 18 | 50 | 2 | 33.33 | 30 | 71.43 | 44 | 61.97 | 13 | 92.86 | 85 | 42.93 |
| **Hormone Induced** | 23 | 20.72 | 47 | 69.12 | 11 | 30.56 | 21 | 52.5 | 3 | 30 | 18 | 50 | 4 | 66.67 | 12 | 28.57 | 27 | 38.03 | 1 | 7.14 | 113 | 57.07 |

**Supplemental Table IV.** Naturally or hormonally induced periods in the last 3 months presented by study group and by site of data collection. Data shown as percentages (%) by study group.
