## Supplementary material for "Clinical profiling of specific diagnostic subgroups of women with chronic pelvic pain": Assessment-tools employed at each study phase based on the domains of interest.

| **Domain** | **Tool** | **Study Component** | **Study Phase** |
| --- | --- | --- | --- |
| Pain: |  |  |  |
| - Clinical history of pelvic pain | EPHect clinical covariates questionnaire (23) | Baseline questionnaire  Follow-up questionnaire | I  II |
| - Intensity | NRS | Baseline questionnaire  Follow-up questionnaire  How are you today? | I  II  III |
| - Nature | i) painDETECT (24)  ii) Short-form McGill Pain Questionnaire 2 (SFMPQ2) (25) | Pain Characteristics | II |
| - Location | i) Fibromyalgia survey scale (26)  ii) Body map | Other symptoms and experiences  How are you today? | II    III |
| - Comorbid pain conditions | i) Diagnoses given  ii) Complex medical symptoms inventory (27)  iii) TMD-pain screening tool (28) | Baseline questionnaire  Follow-up questionnaire  Other symptoms and experiences | I  II    II |
| Gastrointestinal symptoms | i) EPHect clinical covariates questionnaire (23)  ii) The Gastrointestinal Symptom Rating Scale (29) | Baseline questionnaire  Follow-up questionnaire  Other symptoms and experiences | I  II |
| Mood | Hospital Anxiety and Depression Scale (HADS) (30) | Other symptoms and experiences | II |
|  | State anxiety inventory (31) | How are you today? | III |
| Pain Cognitions | Pain Catastrophising Scale (32) | Baseline questionnaire  Follow-up questionnaire  How are you today | I  II    III |
| Personality | Personality Inventory (33) | Other symptoms and experiences | II |
| Somatosensory processing | i) QST (34) on the lower abdomen (test site) and dorsum of the foot (control site)  ii) fMRI with punctate stimuli on the thigh | Physiological testing  fMRI | III  IV |
| Visceral sensitivity | Bladder Sensitivity paradigm (35) | Physiological testing | III |
| Autonomic nervous system function | i) heart rate monitoring before and after pain stimuli | Physiological testing | III |
| HPA axis function | i) 24 hour cortisol profile (saliva)  ii) cortisol before and after pain stimuli (saliva) | Physiological testing | III |
| Endogenous pain modulation | Conditioned pain modulation paradigm (36) | Physiological testing | III |
| Flares in symptoms and triggers | Characteristics of flares in symptoms and associated triggers (37) | Flares questionnaire | II |
| Sleep and fatigue | i) ASCQ-Me v2 Sleep Impact Short Form (38)  ii) Neuro-QOL v1 Fatigue (39) | Other symptoms and experiences | II |
| Medical history | i) EPHect clinical covariates questionnaire  ii) Allergies | Baseline questionnaire  Follow-up questionnaire  Other symptoms and experiences | I  II    II |
| Trauma | Childhood traumatic events scale (40)  Recent traumatic events scale (40) | Other symptoms and experiences | II |
| Treatments | Treatments tried questions with scoring of outcome | Treatments tried questionnaire | II |
| Overall health | EPHect clinical covariates questionnaire | Baseline questionnaire  Follow-up questionnaire | I  II |

**Supplemental Table V.** Assessment-tools employed at each study phase based on the domains of interest.
